# Health Belief, Planned Behavior, or Psychological Antecedents: What predicts COVID-19 Vaccine Hesitancy better among the Bangladeshi Adults?

**DOI:** 10.1101/2021.04.19.21255578

**Authors:** Mohammad Bellal Hossain, Md. Zakiul Alam, Md. Syful Islam, Shafayat Sultan, Md. Mahir Faysal, Sharmin Rima, Md. Anwer Hossain, Abdullah Al Mamun

**Affiliations:** Department of Population Sciences, University of Dhaka, Dhaka, Bangladesh; Department of Population Science, Jatiya Kabi Kazi Nazrul Islam University, Mymensingh; Ovibashi Karmi Unnayan Program (OKUP), Dhaka

## Abstract

**Background:** This study aimed to determine the prevalence and investigate the constellations of psychological determinants of the COVID-19 vaccine hesitancy among the Bangladeshi adult population utilizing the health belief model-HBM (perceived susceptibility to and severity of COVID-19, perceived benefits of and barriers to COVID-19 vaccination, and cues to action), the theory of planned behavior-TPB (attitude toward COVId-19 vaccine, subjective norm, perceived behavioral control, and anticipated regret), and the novel 5C psychological antecedents (confidence, constraints, complacency, calculation, and collective responsibility). We compared the predictability of these theoretical frameworks to see which framework explains the highest variance in COVID-19 vaccine hesitancy.

**Methods:** This study adopted a cross-sectional research design. We collected data from a nationally representative sample of 1497 respondents through both online and face-to-face interviews. We employed multiple linear regression analysis to assess the predictability of each model of COVID-19 vaccine hesitancy.

**Results:** We found a 41.1% prevalence of COVID-19 vaccine hesitancy among our study respondents. After controlling the effects of socio-economic, demographic, and other COVID-19 related covariates, we found that the TPB has the highest predictive power (adjusted R^2^ =0.43), followed by the 5C psychological antecedents of vaccination (adjusted R^2^ =0.32) and the HBM (adjusted R^2^ =0.31) in terms of explaining total variance in the COVID-19 vaccine hesitancy among the adults of Bangladesh.

**Conclusions:** This study provides evidence that theoretical frameworks like the HBM, the TPB, and the 5C psychological antecedents can be used to explore the psychological determinants of vaccine hesitancy, where the TBP has the highest predictability. Our findings can be used to design targeted interventions to reduce vaccine hesitancy and increase vaccine uptake.

**Key Questions:** *What is already known?:*

⍰ There is a global-level insurgence of COVID-19 vaccine hesitancy, where the majority of studies come from western, educated, industrialized, rich, and democratic (WEIRD) countries.
⍰ To date, an online survey found that the prevalence of COVID-19 vaccine hesitancy in Bangladesh was 32.5%.
⍰ Few studies from WEIRD countries have adopted the Health Believe Model and/or the Theory of Planned Behavior to explore the predictors of COVID-19 vaccine hesitancy.

*What are the new findings?:*

⍰ This study found a 41.1% prevalence of COVID-19 vaccine hesitancy among a nationally representative sample of Bangladesh.
⍰ After controlling the effects of demographic, socio-economic, and other COVID-19 related covariates, we found that the TPB has the highest predictive power, followed by the 5C psychological antecedents and the HBM in terms of explaining total variance in the COVID-19 vaccine hesitancy among the adults of Bangladesh.

*What do the new findings imply?:*

⍰ Theoretical frameworks like the HBM, the TPB, and the 5C psychological antecedents can be used to explore the multitude of the psychological determinants of vaccine hesitancy, where the TPB has the highest predictability.
⍰ Findings can be used to design targeted interventions to reduce COVID-19 vaccine hesitancy and increase vaccine demand and uptake.

## INTRODUCTION

Though vaccines, in the form of successful mass immunization programs, have saved millions of lives and improved health and wellbeing across the world ^1^, historically, such successes have constantly been challenged by a minority, yet a significant proportion of ‘vaccine-hesitant individuals and groups for a variety of environmental, cultural, political and psychological reasons ^2–5^. The World Health Organization has identified vaccine hesitancy as one of the top ten global health threats ^6^, where vaccine hesitancy has been defined as a delay in acceptance or refusal of vaccination despite its availability ^5^.

The majority of the studies on vaccine hesitancy comes from the western, educated, industrialized, rich, and democratic (WEIRD) countries ^7–9^. Thus, very little is known about the contexts of developing, low- and middle-income countries ^10^. The world has once again witnessed an insurgence of vaccine hesitancy globally during the pandemic of Coronavirus Disease 2019 (COVID-19)^11^. A systematic review found that many studies reported a COVID-19 vaccine acceptance rate below 60%, ranging from the lowest 23.6% in Kuwait to the highest 97% in Ecuador ^9^.

Studies recommend that 60-75% immunity at the population level would be necessary to halt the Coronavirus transmission and community spread ^12,13^. Thus, the Government of Bangladesh aims to achieve an 80% coverage of the COVID-19 vaccination program ^14^. However, there is an apparent dearth of research on COVID-19 vaccination behavior in Bangladesh. To date, an online survey found that the prevalence of vaccine hesitancy was 32.5% among the adult population of Bangladesh aged 18 years and above, though this study lacks generalizability and was conducted before the vaccine was publicly available ^15^. Research on vaccine hesitancy has received less attention in Bangladesh until the recent pandemic of COVID-19 due to the success stories of expanded programs on immunization^16,17^

Along with quantifying the prevalence of vaccine hesitancy among the population, it is crucial to understand the determinants of the individual decision-making process that result in delay or omission of vaccination ^18,19^. Because vaccine hesitancy reduces vaccine demand and the uptake ^5^, which can impact vaccination coverage, that may ultimately hinder the successful control of the COVID-19 pandemic in Bangladesh. This study, therefore, aimed to determine the prevalence of COVID-19 vaccine hesitancy and investigate the constellations of psychological determinants of vaccine hesitancy among the adult population of Bangladesh, utilizing three of the most widely used theoretical frameworks and models-, the Health Belief Model (HBM) ^20,21^, the Theory of Planned Behavior (TPB) ^22,23^ and the novel 5C psychological antecedents of vaccination ^10,24^.

### Vaccine hesitancy: theories and models to understand vaccination behavior

Vaccine hesitancy is a complex, emerging term in socio-medical literature that recognizes a continuum between total acceptance and outright refusal of some or all vaccines ^5,19^. Hence, vaccine-hesitant individuals comprise a heterogeneous group in the middle of a continuum ranging from the total acceptors of vaccine to the complete refusers ^4^.

The complex nature of motives behind vaccine hesitancy has primarily been explored and analyzed using the epidemiological triad of *environment* (contextual factors), agent (vaccine and disease), and host factors (individual characteristics) ^1,19^. Building on this triad, the WHO Strategic Advisory Group of Experts (SAGE) on immunization drafted a “Model of determinants of vaccine hesitancy” around three key domains: 1) Contextual influences – comprising historical, socio-cultural, environmental, health system/institutional, economic or political factors; 2) Individual and group influences – including influences arising from the personal perception of and attitude towards the vaccine or influences of the social/peer environment; and 3) Vaccine and vaccination-specific issues those are associated with the vaccine characteristics or the vaccination process ^4,5^.

However, in this study, our focus was explicitly directed towards the psychological determinants of vaccine hesitancy ^18,19^, as vaccine hesitancy is predominantly the outcome of the individual decision-making process, which is influenced by individual’s feeling about the vaccination or a particular vaccine, barriers, and enablers to vaccinate ^10,18,19^. Thus, it is crucial to understand which psychological determinants or drivers determine to delay or refusal of the vaccination ^18,19^ so that targeted interventions can be designed to reduce vaccine hesitancy and increase vaccine demand ^5,25^. Therefore, to fulfill that intent, we have adopted two of the most widely used social cognition models-the HBM ^26^ and the TPB ^27^, and the latest framework of 5C psychological antecedents of vaccination^10^ to explore and predict the COVID-19 vaccine hesitancy among adults of Bangladesh.

The HBM is one of the most widely used models in vaccination behavior, particularly in influenza ^28^, swine flu ^22^, ebola ^28^, and hepatitis B ^29^. Few studies have investigated COVID-19 vaccination behavior utilizing the HBM ^20,21,30^. The key argument of HBM is that the likelihood of an individual adopting a particular health behavior (e.g., getting COVID-19 vaccine) is determined by the perceived susceptibility and severity of illness or disease (e.g., COVID-19), along with the belief in the effectiveness of the recommended health behavior (e.g., COVID-19 vaccination) ^26^. The model is comprised of, as applied to COVID-19 and its vaccine, *perceived severity of and perceived susceptibility* to COVID-19, *perceived benefits of and perceived barriers* to getting a COVID-19 vaccine, and *cues to action* which include implicit or explicit incentives or situations that serve to motivate vaccination, such as, information from mass media ^30^.

In contrast, the TPB argues that behavior is driven by the intention to carry out the behavior, which ultimately is determined by an individual’s ‘belief structure’ ^27^. As applied to the context of COVID-19 vaccine, belief structure is comprised of *attitude towards COVID-19 vaccine* (i.e., its perceived necessity, benefit, and effectiveness), subjective norms (i.e., whether significant others support getting a COVID-19 vaccine), and *perceived behavioral control* (i.e., to what extent COVID-19 vaccination is perceived within the individual’s control) ^31^. However, new components are continually being added to the TPB framework to increase its usefulness ^22^. For example, Gallagher and Povey (2006) found that the addition of ‘anticipated regret’ substantially increased the predictive value of TPB in terms of older adults’ intention to get a seasonal influenza vaccination.

A few studies have combined the HBM and the TPB to identify health-related behaviors and explore the multitude of the psychological determinants that influence individual decision-making, such as receiving influenza vaccine among the general public ^22,32^ or in the context of COVID-19 vaccination ^30,31^. Gerund & Shepherd (2012) found that the TPB explained more variance and produced a better model fit than the HBM ^33^.

Finally, Betsch and colleagues (2018) have incorporated and expanded existing vaccination behavior measures and proposed a novel framework of 5C psychological antecedents of vaccination. It includes *confidence* (trust in vaccine effectiveness, safety, and necessity, and the system that delivers it), *complacency* (perceiving the disease as low risk), *constraints* (perceived low vaccine availability, affordability, accessibility, and other barriers to vaccinating), *calculation* (analyzing pros and cons of vaccination), and *collective responsibility* (willingness to take the vaccine for protecting others via herd immunity). Compared with other existing models, the 5C psychological antecedents have explained a greater extent of vaccination variance. However, it has not been tested alongside the HBM and the TPB ^10^.

## METHODS

### Study Design and Data Collection

The study adopted a cross-sectional research design. The calculated sample size was 1635, where the Z-score for 95% confidence interval was 1.96, prevalence (p) of COVID-19 vaccine hesitancy from a previous study was 0.33^15^, the margin of error (e) was 0.03, design effect (Deff) for sampling variation was 1.6, and a non-response rate (NR) was 10%. However, 112 respondents did not consent to participate in the survey, while another 26 respondents did not know about the COVID-19 vaccine. After excluding them, the final sample stood 1497 for the analysis.

We collected data from all eight administrative divisions of Bangladesh using probability-proportionate to each division’s population size. Both online and face-to-face interviews were conducted to collect data. Data were collected using the online platform Google form from one-third of the respondents. A link to the survey questionnaire was created and sent to the prospective respondents via e-mail, WhatsApp, and Facebook messenger. All the respondents to whom the survey link was sent were requested to share the link in their network to reach more people. The research team members circulated the survey link in their respective professional and social networks. The online link was valid for three days. After that, the collected data were downloaded, and divisional distribution was assessed. We then collected data for the remaining sample size of each division using face-to-face interviews. For this, we randomly selected two districts from each division. Within each district, the sample was distributed proportionately according to the rural-urban distribution of its population. Then, convenience sampling was adopted to accomplish face-to-face interviews from the population-based households.

The respondent selection criteria for the face-to-face interview were adult people of 18 years and above living in Bangladesh and knowing about the COVID-19 vaccine. Reading and writing ability and using the Internet were added criteria for selecting the online survey respondents. The data is now available in the Mendeley open research data repository ^34^.

## Measures

### Outcome Variable: COVID-19 Vaccine Hesitancy

We used two 6-points Likert-type items to measure COVID-19 vaccine hesitancy among the respondents: (a) *If you get the chance of getting a COVID 19 vaccine for free, what will you do?* (with the response of 1= *Surely, I will take it*; 2= *Probably I will take it*; 3= *I will delay taking it*; 4= *I am not sure what I will do*; 5= *Probably I will not take it*; 6= *Surely, I will not take it*), and (b) *If your family or friends think of getting COVID 19 vaccine, what will you do?* (with the response of 1= *Strongly encourage them*; 2=*Encourage them*; 3=*Ask them to delay getting the vaccine*; 4=I *will not say anything about it*; 5=*Discourage them to take vaccine*; 6=*Forbid them to take vaccine*). We combined these two items and calculated the level of COVID-19 vaccine hesitancy, where a higher score indicated a higher level of hesitancy toward the COVID-19 vaccine. The theoretical total score ranges from 2 to 12. The reliability analysis of the scale demonstrates an excellent internal consistency (Cronbach Alpha=0.833).

### Predictor Variables

#### The HBM constructs

The HBM constructs consisted of the following components: perceived susceptibility (included two items, α=0.657), perceived severity (included two items, α=0.612), perceived benefits (included three items, α=0.841), perceived barriers (included five items, α=0.735), and cues to action (Table 1). These items were measured on a 5-point Likert scale (1= Strongly Disagree to 5= Strong Agree). Cues to action included self-reported COVID-19 positive status for self and family members, and sources of vaccine-related knowledge (social media/ online news portals/blog, and print media).

**Table 1:** Items used to measure HBM, TPB, and 5C psychological antecedents.

| Statements | Cronbach $\alpha$ |
| --- | --- |
| **The Health Belief Model** <sup>26</sup> |  |
| **Perceived Susceptibility** | **0.657** |
| I am worried about the likelihood of getting infected by COVID-19 |  |
| I am at high risk of COVID-19 because of my health conditions |  |
| **Perceived Severity** | **0.612** |
| I will be very sick if I get infected by COVID-19 |  |
| I am very concerned that I could die from COVID-19 |  |
| **Perceived Benefits** | **0.841** |
| I think vaccination is good because it will make me less worried about COVID-19 |  |
| I believe vaccination will decrease my risk of getting infected by COVID-19 |  |
| I think the complications of COVID-19 will decrease if I get vaccinated and then get infected with the Coronavirus. |  |
| **Perceived Barriers** | **0.735** |
| I am worried that the possible side effects of the COVID-19 vaccination would interfere with my usual activities |  |
| I am concerned about the efficacy of the COVID-19 vaccine |  |
| I have a concern that I may receive faulty/fake COVID-19 vaccine |  |
| It concerns me that the development of a COVID-19 vaccine is too rushed to test its safety properly |  |
| I am concerned about the long-term side effects of the COVID-19 vaccination |  |
| **Cues to Action** | **NA** |
| Respondents got infected with Coronavirus |  |
| A family member got Infected with Coronavirus |  |
| Social media (e.g., Facebook) or online news portals/blog as a source of knowledge about the COVID-19 vaccine |  |
| Printed newspaper as a source of knowledge about COVID-19 vaccine |  |
| **The Theory of Planned Behavior** <sup>27</sup> |  |
| **Attitude toward Vaccine** | **0.739** |
| I think the COVID-19 vaccine probably will not work |  |
| I do not trust the COVID-19 vaccine |  |
| I think the COVID-19 vaccine is unnecessary |  |
| I think it is not important to get a vaccine to protect people from the COVID-19 |  |
| I do not need a COVID-19 vaccine because I am healthy and at low risk for infection |  |
| I do not need a COVID-19 vaccine because even if I get infected, I will not become seriously ill |  |
| **Subjective Norm** | NA |
| I believe my family members will support me to get vaccinated against COVID-19 |  |
| **Perceived Behavioral Control** | NA |
| If I want, I can register for COVID 19 vaccination |  |
| **Anticipated Regret** | NA |
| If I do not get a COVID-19 vaccine and end up getting Coronavirus, I will regret not getting the vaccination |  |
| **The 5C Psychological Antecedents of Vaccination <sup>10</sup>** |  |
| **Confidence** | **0.808** |
| I am completely confident that COVID 19 vaccines are safe |  |
| I am completely confident that COVID 19 vaccines are effective |  |
| Regarding COVID 19 vaccines, I am confident that public authorities decide in the best interest of the community |  |
| **Constraints** | NA |
| Everyday work stress may prevent me from getting vaccinated |  |
| **Complacency** | **0.753** |
| I think it is unnecessary to receive vaccinations as it cannot prevent COVID-19 |  |
| I believe my immune system is powerful; it will protect me from COVID-19 |  |
| I believe COVID-19 is not much a severe disease that I should get vaccinated against it |  |
| **Calculation** | **0.909** |
| When I think about getting vaccinated against COVID 19, I weigh the benefits and risks to make the best decision possible |  |
| When I think about getting vaccinated against COVID 19, I will first consider whether it is effective or not |  |
| Before I get COVID-19 vaccinated, I need to know about this vaccine in details |  |
| **Collective Responsibility** | **0.612** |
| I will take COVID 19 vaccine because, in that way, I can protect people with a weaker immune system |  |
| I think vaccination against COVID 19 is a collective action to prevent the spread of diseases |  |

#### The TPB constructs

The TPB constructs consisted of four domains: attitude toward vaccine (including six items, α=0.739), subjective norm, perceived behavioral control, and anticipated regret (Table 1). Each item of the four domains was assessed on a 5-point scale (1 = *strongly disagree* to 5 = *strongly agree*). The mean score of the items under attitude toward vaccine component was calculated, with a higher average score indicating more negative attitude toward COVID-19 vaccine.

#### The 5C psychological antecedents of vaccination

The 5C psychological antecedents of vaccination consisted of 5 antecedents: confidence (included three items, α=0.808), complacency (included three items, α=0.753), constraints (included a single item), the calculation (included three items, α=0.909), and collective responsibility (included two items, α=0.612) (Table 1). Each item of the five antecedent domains was assessed on a 5-point scale (1 = *strongly disagree*; 5 = *strongly agree*). Except for the constraints domain, mean scores of items under each domain were computed, with a higher average score indicating the corresponding domain’s stronger agreement.

### Other Covariates

We also collected data on the following independent variables: age, sex, religion, marital status, educational attainment, place of residence, geographic region, occupation, number of family members, household income, knowledge about COVID-19 vaccine, knowledge about vaccination process, and behavioral practice to prevent COVID-19 (Table 2).

**Table 2.** Sample characteristics of the respondents.

| Variables | Estimates, n (%) |
| --- | --- |
| **Age in years** |  |
| 18-24 | 432 (28.9) |
| 25-30 | 362 (24.2) |
| 31-39 | 254 (17.0) |
| 40-49 | 236 (15.8) |
| 50+ | 213 (14.2) |
| Average Age, Mean (SD) | 33.7 (12.9) |
| **Sex** |  |
| Women | 692 (46.2) |
| Men | 805 (53.8) |
| **Religion** |  |
| Others | 196 (13.1) |
| Muslim | 1301 (86.9) |
| **Marital status** |  |
| Never-married | 575 (38.4) |
| Ever-married | 922 (61.6) |
| **Education** |  |
| No education | 129 (8.6) |
| Primary | 179 (12.0) |
| Secondary and higher secondary | 448 (29.9) |
| Graduate | 400 (26.7) |
| Masters and MPhil/PhD | 341 (22.8) |
| **Place of residence** |  |
| Rural area | 963 (64.3) |
| Urban area (other than city corporation) | 179 (12.0) |
| City Corporation | 355 (23.7) |
| **Administrative division** |  |
| Barishal | 114 (7.6) |
| Chattogram | 253 (16.9) |
| Dhaka | 478 (31.9) |
| Khulna | 137 (9.2) |
| Mymensingh | 108 (7.2) |
| Rajshahi | 180 (12.0) |
| Rangpur | 114 (7.6) |
| Sylhet | 113 (7.5) |
| **Occupation** |  |
| Government, private, & NGO sector job | 202 (13.5) |
| Professional (teacher, engineer, lawyer, doctor, nurse, paramedics, pharmacist) | 277 (18.5) |
| Housewife | 348 (23.2) |
| Students and unemployed | 473 (31.6) |
| Agriculture and day Laborer | 102 (6.8) |
| Others | 95 (6.3) |
| **Household members, Mean (SD)** | 4.9 (1.98) |
| **Household income in BDT, Mean (SD)** | 37627.2<br>(81295.9) |
| **Knowledge about COVID-19 vaccine, Mean (SD)** | 11.4 (2.2) |
| **Knowledge about vaccination process, Mean (SD)** | 2.8 (2.0) |
| **Behavioral practice to prevent COVID-19, Mean (SD)** | 8.8 (2.7) |
| **Total** | **1497 (100.0)** |

#### Knowledge about COVID-19 vaccine

We assessed the knowledge related to the COVID-19 vaccine using four Likert-type items: *COVID-19 vaccine is safe for pregnant women, COVID-19 vaccine is safe for children under 18 years old, COVID-19 vaccine have a very mild side effect, and side effects COVID-19 vaccine do not last longer than 2 days*. The response options against these items were “strongly disagree = 1”, “disagree = 2”, “neither agree nor disagree = 3”, “agree = 4”, and “strongly agree = 5”. The total score of these items ranged between 4 to 20, with a higher score indicating higher knowledge. Reliability analysis of the scale suggested an acceptable internal consistency (Cronbach alpha, α=0.643).

#### Knowledge about vaccination process

Knowledge about the COVID-19 vaccination process was measured by the following six binary response (yes=1, no=0) questions: (1) do you know that you need to consult with a doctor to receive the COVID-19 vaccination?; (2) do you know that you need to register online to receive the COVID-19 vaccination?; (3) do you know that you can receive the COVID-19 vaccination only from selected health facilities?; (4) do you know that you cannot receive the COVID-19 vaccination from a pharmacy?; (5) do you know that health care workers will not provide the COVID-19 vaccine at your doorstep?; and (6) do you know the correct doses of the COVID 19 vaccine? The reliability analysis showed good internal consistency among these six questions (α=0.765). The total score ranged between 0 to 6, with a higher score indicating better knowledge about the COVID-19 vaccination process.

#### Behavioral practice to prevent COVID-19

The level of preventive behavioral practices related to COVID-19 was measured using three Likert-type items: *I avoid crowds as much as possible to prevent my risk of getting COVID-19, I always wear a mask when I am outside of my home or around other people, and I am conscious about using sanitizer or hand wash*. The response options against these items were “never = 1”, “sometimes = 2”, “often = 3”, and “regularly = 4”. The total score ranged between 3 to 12, with a higher score indicating a higher level of preventive practices. The reliability analysis showed good internal consistency in this scale (α=0.857).

### Statistical Analysis

We employed multiple linear regression analysis to assess the selected model’s predictability to COVID-19 vaccine hesitancy after checking the assumptions and multicollinearity. We produced three models. In each model, significant demographic, socio-economic, knowledge, and practice of COVID-19 related variables were controlled to predict the used model. Akaike Information Criterion (AIC), Amemiya Prediction Criterion (APC), Mallows’s Prediction Criterion (MPC), and Schwarz Bayesian Criterion (SBC) were used to assess model performance.

### Patient and Public Involvement

No public or patients were involved in the process of this research.

### Human Subject’s Protections

The present study was carried out following the Declaration of Helsinki. The respondents were informed about the aims, objectives, potential scopes, and implications of this study’s findings and were requested to participate voluntarily.

## RESULTS

### Background Characteristics of the Respondents

The demographic, socio-economic, and other background characteristics of the respondents are summarized in Table 2. The respondents’ average age was 33.7 years, with a standard deviation (SD) of 12.9. The highest proportion (28.9%) of respondents was from 18–24 years. About 47% of the respondents were women, while most of the respondents (86.9%) were Muslim. About two-thirds of the respondents (61.6%) were married, while 20.6% of the respondents had less than a secondary education level. About two-thirds of the respondents (64.3%) were from rural areas, while 31.9% were from Dhaka Division. One-third (31.6%) of the respondents were students and unemployed. The mean number of household members was 4.9, while the mean household income was BDT 37627 (Table 2).

### Prevalence of COVID-19 Vaccine Hesitancy

Nearly 43% of respondents reported that they would surely take the COVID-19 vaccine if it were available for free, whereas another 17.7% of respondents would probably take it. Thus, a total of 60.6% of the respondents were potential COVID-19 vaccine acceptors. However, 39.4% of respondents were found vaccine-hesitant, where nearly 7% were complete refusers of the COVID-19 vaccine. A similar level of COVID-19 vaccine hesitancy was found among the respondents when asked about their opinion about their family members or relatives’ hypothetical vaccination decisions. A total of 60.3% of the respondents mentioned that they would strongly encourage or encourage their family members or relatives if they decided to take the COVID-19 vaccine. Thus, rest 39.7% of respondents were vaccine-hesitant in terms of their family members and relatives’ decision to vaccinate, where 3% would forbid them to take the COVID-19 vaccine. The mean score of the hesitancy scale was 4.93 with an SD of 2.68. The accuracy of the hesitancy scale was 41.1% (4.93/12*100), indicating that 41.1% of the respondents had hesitancy to accept the COVID-19 vaccine.

### Predictors of COVID-19 Vaccine Hesitancy in Bangladesh

After controlling the effects of demographic, socio-economic, and other COVID-19 related covariates, our first model shows that the HBM constructs explained 31% of the variance in COVID-19 vaccine hesitancy (adjusted R^2^=0.31) (Table 3). Among the HBM constructs, perceived susceptibility, perceived severity, perceived benefits, and perceived barriers were the significant predictors of COVID-19 vaccine hesitancy. An increase in perceived susceptibility of COVID-19 tended to reduce vaccine hesitancy (β=−0.06, p<0.05). Similarly, increased perceived severity of COVID-19 also reduced vaccine hesitancy (β=−0.11, p<0.01). However, among the HBM constructs, perceived benefits had the largest standardized co-efficient. In other words, a one-unit increase in perceived benefits of getting the COVID-19 vaccine reduced the vaccine hesitancy by 0.33 units (β=−0.33, p<0.01). On the other hand, perceived barriers to getting the COVID-19 vaccine tended to increase vaccine hesitancy (β= 0.30, p<0.01). However, the cues to action construct were found insignificant in predicting COVID-19 vaccine hesitancy, except the component ‘Social media (e.g., Facebook) or online news portals/blog as a source of knowledge about the COVID-19 vaccine’. In other words, respondents who heard about the COVID-19 vaccine from social media (e.g., Facebook) or online news portals were less vaccine-hesitant (β= −0.05, p<0.05).

**Table 3.**
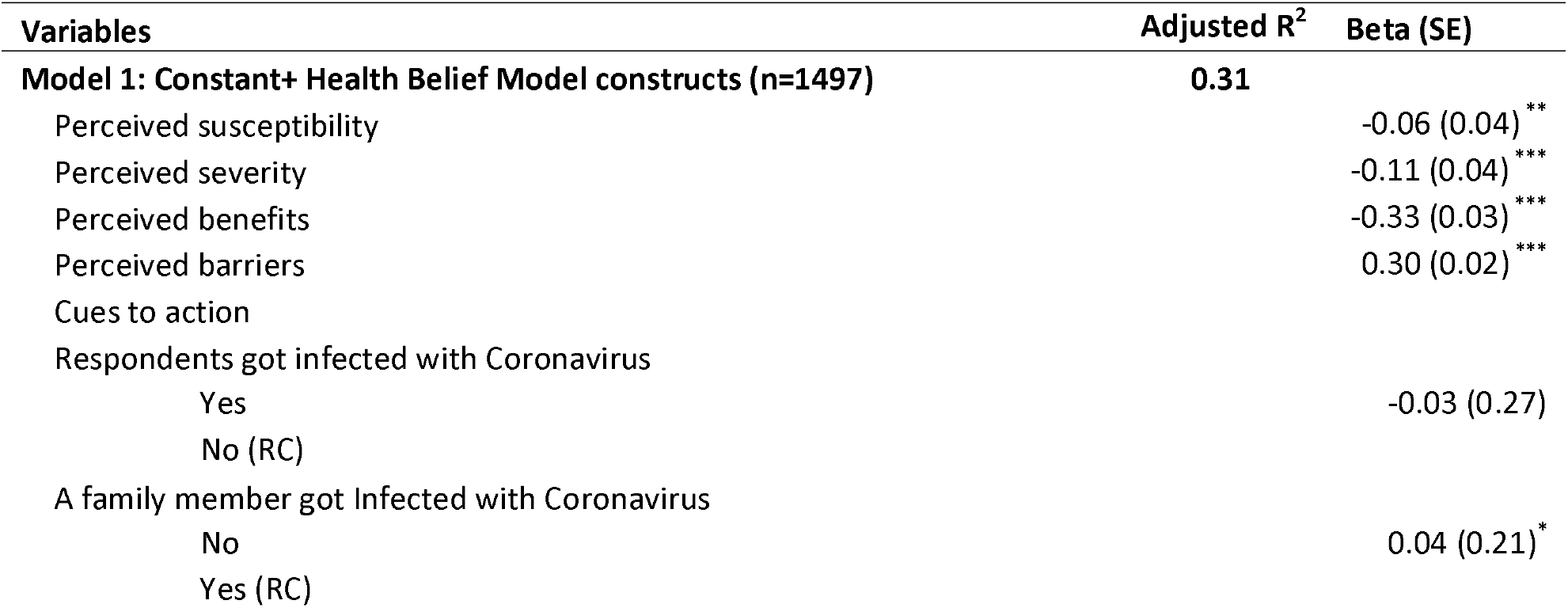
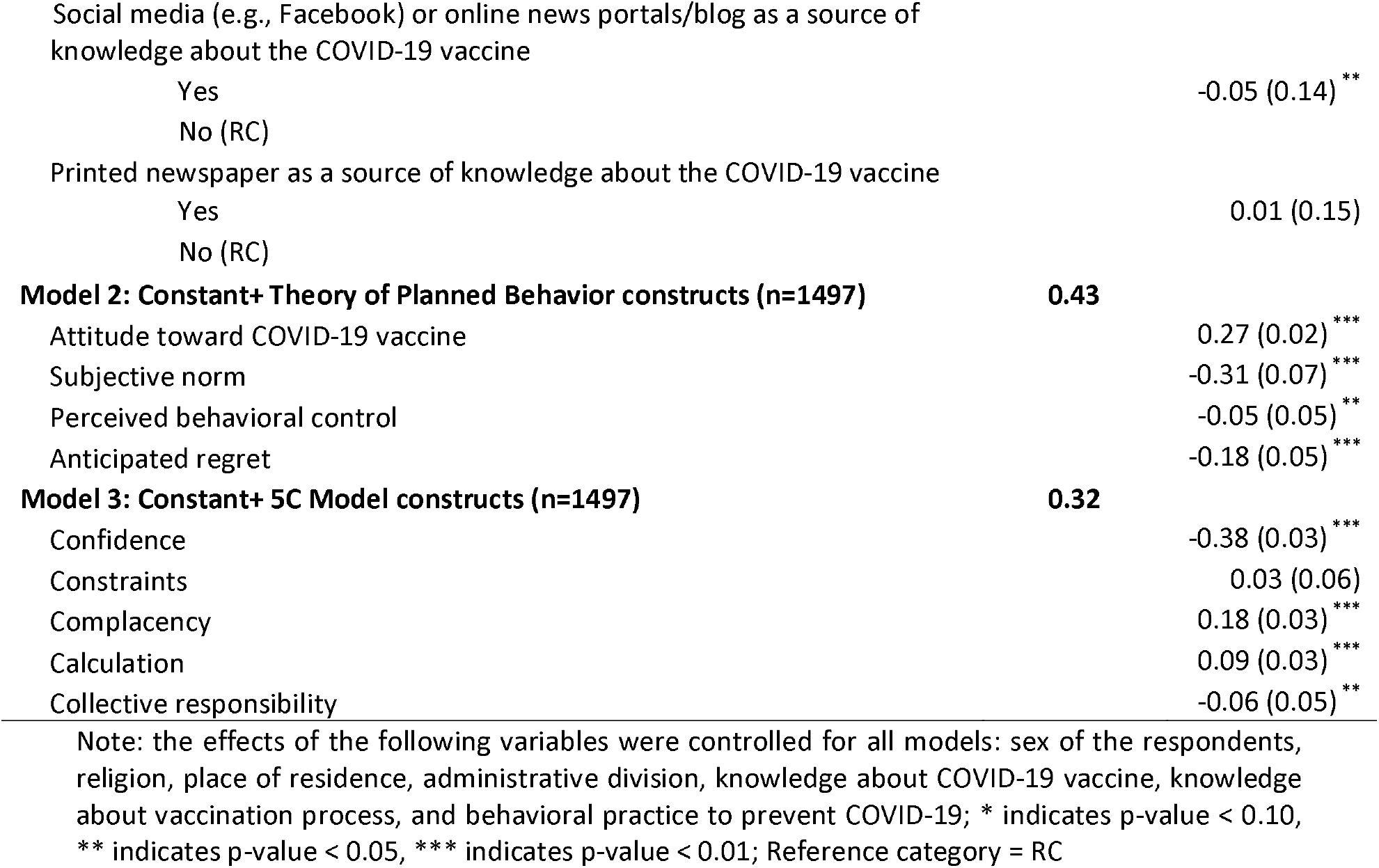
Multiple linear regression analysis to predict COVID-19 vaccine hesitancy.

The second model shows that 43% of the variance in COVID-19 vaccine hesitancy was explained by the TPB (adjusted R^2^=0.43) (Table 3). All four components of the TPB were the significant predictors of COVID-19 vaccine hesitancy. Respondents who had a more negative attitude toward the COVID-19 vaccine were more vaccine-hesitant (β=0.27, p<0.01). Vaccine hesitancy tended to decrease with the increase of familial support in favor of vaccination regarding the TPB’s subjective norm (β=-0.31, p<0.01). However, in terms of perceived behavioral control, respondents who mentioned registering for COVID-19 vaccination was within their control were less vaccine-hesitant (β=-0.05, p<0.05). Finally, an increase in anticipated regret among the respondents resulted in reduced vaccine hesitancy (β=-0.18, p<0.01).

The third model included the 5C psychological antecedents, which explained 32% of the variance in COVID-19 vaccine hesitancy (adjusted R^2^=0.32) (Table 3). An increase in COVID-19 vaccine confidence tended to decrease vaccine hesitancy (β=-0.38, p<0.05). The more complacent individuals had a more vaccine hesitancy (β=0.18, p<0.01). Similarly, respondents who were more calculative about the pros and cons of getting vaccinated or needed more information about the vaccine before getting vaccinated were significantly more COVID-19 vaccine-hesitant (β=0.09, p<0.01). Finally, respondents who had a sense of collective responsibility to vaccinate against COVID-19 were significantly less vaccine-hesitant (β=-0.06, p<0.05).

Table 4 presents the model summary and the selection criteria to assess model performance. For each of the three multiple linear regression models to identify psychological determinants of COVID-19 vaccine hesitancy, we compared AIC, APC, MPC, SBC to select the best fitting model. The model with the smallest AIC, APC, MPC, SBC values was the best-fitted model. According to these indicators, the TPB constructs (Model 2) were the best-fitted model (Table 4).

**Table 4.** Model Summary and Selection Criteria.

| Model Summary | Model 1: HBM | Model 2: TPB | Model 3: 5c |
| --- | --- | --- | --- |
| n | 1497 | 1497 | 1497 |
| R | 0.56 | 0.66 | 0.57 |
| R<sup>2</sup> | 0.32 | 0.43 | 0.33 |
| Adjusted R<sup>2</sup> | 0.31 | 0.43 | 0.32 |
| F Change | 108.2 | 205.09 | 92.69 |
| df | 4, 1474 | 4, 1478 | 5, 1477 |
| Significance of F Change | <.001 | <.001 | <.001 |
| **Selection Criteria** |  |  |  |
| Akaike Information Criterion (AIC) | 2427.1 | 2144.6 | 2399.0 |
| Amemiya Prediction Criterion (APC) | 0.70 | .58 | 0.69 |
| Mallows' Prediction Criterion (MPC) | 23.0 | 19.0 | 20.0 |
| Schwarz Bayesian Criterion (SBC) | 2549.2 | 2245.5 | 2505.23 |

## DISCUSSION

Given the dearth of research on COVID-19 vaccination behavior in Bangladesh, this study aimed to determine the prevalence of COVID-19 vaccine hesitancy and explore its psychological determinants among Bangladeshi adults. To fulfill that intent, we have utilized three of the most widely used tools in the field of vaccination behavior-the HBM, the TPB, and the novel 5C psychological antecedents of vaccination, and compared their predictability in terms of predicting the COVID-19 vaccine hesitancy.

We found a 41.1% prevalence of the COVID-19 vaccine hesitancy among our study respondents, a significantly higher estimate than found by Ali and colleagues (2021) (32.5%). This higher estimation may, in part, be explained by the fact that Ali and colleagues (2021) conducted an online survey which may biasedly present the view of ‘Netigens’ of Bangladesh ^35^. In contrast, in our study, data were collected through online and face-to-face interviews from a nationally representative sample covering all eight administrative divisions. In that sense, our study findings provide a more accurate estimate of COVID-19 vaccine hesitancy that is generalizable in the context of the adult population living in Bangladesh.

However, as per the second objective of this study, we found that after controlling the effects of socio-economic and demographic variables, level of knowledge related to COVID-19, its vaccine and vaccination process, and level of preventive practices towards COVID-19, the TPB has the highest predictive power (adjusted R^2^=0.43), followed by the 5C model (adjusted R^2^=0.32) and the HBM (adjusted R^2^=0.31) in terms of explaining total variance in COVID-19 vaccine hesitancy among the adults of Bangladesh. This finding is particularly unique because, to date, no study compared the predictability of these three behavioral frameworks or models in terms of predicting vaccine hesitancy, especially in Bangladesh. Moreover, given the newness of the 5C scale ^10^, the available literature is still scanty that tests this model’s predictive validity ^24,36^. However, various other studies that adopted only the HBM and the TPB validate our findings that TPB constructs are a better predictor of vaccination behavior ^23^, specifically in the intention to vaccinate against swine flu ^22^ human papillomavirus ^33^ or in the context of COVID-19 vaccination ^20,30,31^.

According to the findings of the HBM constructs, an increase in perceived benefits of the COVID-19 vaccine, along with increasing perceived severity of and perceived susceptibility to COVID-19, significantly reduced the vaccine hesitancy. On the other hand, an increase in perceived barriers to getting vaccinated acted as a significant vaccine hesitancy promoter. These findings essentially correspond to other studies related to influenza vaccination ^37^, though Lin et al. (2020) found perceived susceptibility was not a significant predictor of the COVID-19 vaccine hesitancy in China. In terms of cues to action, though COVID-19 infection status (of both self or family members) were non-significant predictors at a 5 percent level of significance. We found that social media (e.g., Facebook) or online news portals as the source of information about the COVID-19 vaccine was the significant predictor, and respondents who heard about the COVID-19 vaccine from social media (e.g., Facebook) or online news portals or blogs were less hesitant. Taken together, these findings suggest that imparting adequate and proper information about the COVID-19 vaccine to the public, along with solid evidence of the safety, efficacy, and benefits of the COVID-19 vaccine, can be a crucial strategy to reduce vaccine hesitancy and increase its demand and actual uptake. In that case, social media and online news portals may act as more effective means than printed newspapers to disseminate COVID-19 related information, as found in our study.

According to the findings of the TPB constructs, an increase of negative attitude towards the COVID-19 vaccine and a decrease in perceived behavioral control significantly increased the COVID-19 vaccine hesitancy. Other studies support these findings ^23,30^, though perceived behavioral control was found non-significant predictor in other contexts ^22,38,39^. However, subjective norms in the form of family members’ support for having COVID-19 vaccination significantly reduced vaccine hesitancy, corresponding to other studies ^23,30^. Consistent with earlier researches ^22,40,41^, anticipated regret was a significant predictor of COVID-19 vaccine hesitancy. This finding implies that an intervention to increase the COVID-19 vaccination uptake should circulate the message that it is better to get vaccinated than regret later. Alternative measures should be devised to reduce the barriers related to COVID-19 vaccination, such as online registration to get the COVID-19 vaccine. This provision is inconvenient, especially for older persons, people living in rural areas, and those who do not have internet access ^42^.

Finally, according to the 5C model, more substantial confidence and higher collective responsibility were significantly associated with reduced COVID-19 vaccine hesitancy, whereas increased complacency and calculation significantly increased the vaccine hesitancy. These findings are supported by other studies in both COVID-19 and other contexts ^24,36^, though calculation and constraints were non-significant predictors of COVID-19 vaccine hesitancy among the nurses in Hong Kong ^24^. In our study, the constraint related to the COVID-19 vaccination was a non-significant predictor of COVID-19 vaccine hesitancy. These findings suggest that public confidence in the vaccine and the health system that delivers the vaccination service are crucial. Widespread misinformation, conspiracy beliefs, and superstitions regarding the COVID-19 vaccine and its potential health hazards have been found to diminish public trust ^43^ that need to be addressed through proper communication. Extensive information searching about the subjective utility of vaccination, as evidenced in earlier studies ^18,19,44^, might have resulted in more vaccine hesitancy as more calculation increased COVID-19 vaccine hesitancy among the respondents, along with complacency. These findings warrant the urgency of re-iterating the risk communication and health benefits message of getting vaccinated to mass people.

### Strengths and Limitations of the Study

This study is the first to explore the COVID-19 vaccine hesitancy among Bangladeshi adults, adopting a large and diversified representative sample. Moreover, this study also explored many psychological antecedents of the COVID-19 vaccine hesitancy utilizing three of the most widely used theoretical tools-the HBM, the TPB, and the 5C psychological antecedents, and used multivariate modeling to identify the most salient predictors. Therefore, this study’s findings can help design targeted intervention to reduce the COVID-19 vaccine hesitancy, which will help the Government of Bangladesh attain the target of 80% vaccination coverage for the COVID-19 vaccine. Another strength of the study is that our study provides the findings on the prevalence of the COVID-19 vaccine hesitancy and its predictors while the COVID-19 vaccine was publicly available in Bangladesh. Finally, in terms of theoretical contribution of the study in the field of vaccination behavior, this study contributes evidence from a non-WEIRD country that, at one hand, assess the predictive validity of the novel 5C psychological antecedents of vaccination^10^ and, on the other hand, validates the theoretical supremacy of the TPB over the HBM and the 5C model in predicting the COVID-19 vaccine hesitancy among the adult population in Bangladesh. However, our study also has some limitations. This study could not use probability sampling completely. We tried to draw our sample following the national population distribution in terms of age, sex, residence, region, marital status, and religion. However, the distribution of education among the respondents is not comparable to national data. Moreover, this study collected self-reported data that may suffer from reporting bias. Finally, this research used a cross-sectional study design which can not establish causality.

## CONCLUSION

This study provides evidence that theoretical frameworks like the TPB, the HBM, and the 5C psychological antecedents can explore the psychological determinants that influence a person’s vaccination decision-making process. Among the frameworks of determinants, the TPB has the highest predictive power in determining the vaccination decision. These findings can be used to craft targeted interventions to reduce vaccine hesitancy and increase vaccine uptake. Thus, this study’s findings will steer Bangladesh’s vaccination campaign and those alike to reach the targeted coverage of the COVID-19 vaccination program and, thereby, paving the way for successfully eradicating the never-ending pandemic of COVID-19.

## Data Availability

The data is now available in the Mendeley open research data repository.

http://dx.doi.org/10.17632/jzvbvvknkv.1

## Acknowledgments

The authors would like to thank the respondents of this study for their valuable time and inputs. We would also like to convey our thanks to selected students of the Department of Population Sciences, the University of Dhaka, for their contribution in conducting field-level data collection amidst this challenging time of the COVID-19 pandemic.

## Contributors

MBH conceptualized the study. All the authors designed the study and collected the data. MBH, MAH, and MZA analyzed and interpreted the data and drafted the manuscript. MSI, SS, MMF, SR, and AAM revised the manuscript critically for valuable intellectual content and approved the final version to be published. All authors remained in agreement to be accountable for all aspects of the work.

## Funding

This study received no funding or whatsoever from any organization or donor agency.

## Competing interests

The authors declared no conflicts of interest.

## Ethical Approval

The study was approved by the Bangladesh Medical Research Council (BMRC).

## Data availability statement

Data associated with this study has been deposited at Mendeley at http://dx.doi.org/10.17632/jzvbvvknkv.1

## Notes

### Competing Interest Statement

The authors have declared no competing interest.

### Funding Statement

No funding was received for this study.

### Author Declarations

The National Research Ethics Committee of the Bangladesh Medical Research Council (BMRC) has approved this study.

